# Perceptions about the Use of Virtual Assistants for Seeking Health Information among Caregivers of Young Childhood Cancer Survivors

**DOI:** 10.1101/2024.08.28.24312737

**Authors:** Emre Sezgin, Daniel I. Jackson, Kate Kaufman, Micah Skeens, Cynthia A. Gerhardt, Emily L. Moscato

## Abstract

**Purpose:** This study examined the perceptions of caregivers of young childhood cancer survivors (YCCS) regarding the use of virtual assistant (VA) technology for health information seeking and care management. The study aim was to understand how VAs can support caregivers, especially those from underserved communities, in navigating health information related to cancer survivorship.

**Methods:** A qualitative study design was employed, involving semi-structured interviews and focus groups with ten caregivers of YCCS from metropolitan, rural and Appalachian regions, recruited from a large pediatric academic medical center in the Midwest. A web-based VA prototype was tested with caregivers, who provided feedback on its usability, utility, and feasibility. Data were analyzed using thematic analysis to identify key themes related to caregivers’ interactions with and perceptions of the VA technology.

**Results:** We identified four major themes: Interface and Interaction, User Experience, Content Relevance, and Trust. Caregivers expressed preferences for multimodal interactions, emphasized the need for accurate and relevant health information, and highlighted the importance of trust and confidentiality. The VA was perceived as a valuable tool for quickly accessing information, reducing the cognitive and emotional burden on caregivers. VAs were perceived to provide tailored support for managing specific health needs of YCCS.

**Conclusions:** VAs hold promise as a support tool for caregivers of YCCS, particularly in underserved communities. By offering personalized, reliable, and easily accessible information, VAs were perceived to support caregivers to manage health conditions and ease the caregiving tasks.

## Introduction

There are over half a million survivors of pediatric cancer in the United States [1, 2]. The complexity of pediatric cancer treatment necessitates that caregivers, often parents, close family members, or legal guardians, navigate an intricate web of information related to diagnosis, treatment options, and side effects [3]. Caregivers receive cancer-related information during the acute treatment phase, as well as in survivorship, when over two-thirds of survivors are at risk of developing one or more chronic health problem, known as late effects [4]. The high cognitive and emotional burden of navigating this process makes it imperative to investigate new avenues of tailored informational support for these caregivers [5, 6].

Young childhood cancer survivors (YCCS), who are diagnosed and treated in early childhood (i.e., less than age 7 years), face the highest risk for late effects given their treatment during a highly sensitive developmental period, particularly for neurodevelopmental late effects [7–9]. Risks are elevated in the context of invasive treatments such as intrathecal chemotherapy and cranial directed radiation [10].

Neurodevelopmental late effects after cancer may include developmental delays (e.g., speech, motor, cognitive), neurocognitive deficits (e.g., memory, attention, executive functioning, and processing speed), and psychosocial difficulties [11]. Thus, caregivers of YCCS must navigate the additional complexity of locating resources related to the diagnosis and treatment of neurodevelopmental conditions and identifying providers with expertise in treating children with medical complexity [7, 12].

Resources to mitigate these neurodevelopmental late effects, such as state-based early intervention and home visiting programs, school liaison support, and rehabilitative therapies (e.g., speech, physical, occupational therapies) may be beneficial, especially when delays are identified and treated early [13]. Still, the availability and quality of these services may vary by geographic location. Specifically, healthcare disparities are known to be more pronounced in rural and Appalachian communities due to factors like geographical isolation, widespread poverty, lack of educational resources, and limited access to specialized medical care [14–16]. Additionally, the extent to which caregivers seek these resources and the use digital tools to identify quality providers in their area remains under-investigated [17].

Existing studies on online health information-seeking behavior reveal that caregivers often prefer personalized, easily accessible, and reliable sources of information [18, 19]. Today, virtual assistants (VAs) such as Amazon’s Alexa, Google Assistant, and Apple’s Siri have become ubiquitous platforms that facilitate various forms of user-technology interaction, from simple tasks like setting reminders to complex ones like accessing health information and care management [20–24]. These VAs have become an integral part of modern information-seeking behavior, with studies indicating a growing preference for voice-based interfaces over traditional textual search in various demographics [25–27]. Notably, with recent advancements in large language models, the capabilities of virtual assistants show improved comprehension and responses [28, 29].

Given the capabilities of VAs in providing information that is not only quick but also personalized through machine learning algorithms [30–33], they may offer a promising solution to the challenges faced by caregivers of YCCS. Caregivers are likely to benefit most if they hold one or more marginalized identities (e.g., racial/ethnic identities, low socioeconomic status) or live in underserved areas (e.g., rural or Appalachian), where resources may be scarce or nonexistent. Yet, we lack empirical data that directly address the adoption, feasibility, and utility of VAs across each of these unique contexts. In particular, the rural and Appalachian communities have been underrepresented in existing research, leading to an incomplete understanding of how these technological tools might be leveraged to meet the unique needs of more isolated populations [34]. Therefore, there is a need to bridge this gap in the literature by investigating perceptions about how VAs could be effectively utilized as an information-sharing tool among caregivers of YCCS, especially amongst those in underserved communities.

The primary aim of this study is to examine the perception of caregivers of YCCS, including those from underserved communities, about VA technology use for health information seeking and care management. Given the potential of VAs to deliver complex, multidimensional information, including that related to neurodevelopmental conditions [35], this study seeks to fill a vital gap in our understanding of how VAs could support caregivers facing these multifaceted challenges and aid in identification and access to early intervention. Furthermore, this study intends to contribute to our broader understanding of how emerging technologies can be adapted to serve marginalized populations, thereby addressing healthcare disparities through targeted, accessible, and high-quality information dissemination.

## Methods

This study used a qualitative approach to understand the perception of caregivers, further elaborating challenges and opportunities associated with the use of Virtual Assistants (VAs) for health information-seeking and care management. The research methodology was informed by constructivist paradigms that acknowledge the subjective experiences of caregivers [36].

### Virtual Assistant Prototype

A web-based virtual assistant was created using a conversational agent platform, Voiceflow [37]. The VA was designed with 3 components; Request, Delivery, and Continuation to simulate naturalistic conversation. (1) Request: The prototype initiated the conversation by requesting a description or question about a topic of interest. The VA was designed to use button responses, voice commands, or free text.

This feature was to demonstrate VA capabilities and provided options to caregivers. (2) Delivery: The VA would respond with relevant information using the knowledge base. This knowledge base included a number of documents referencing from reports by cancer advocacy organizations [38, 39], government policy documents [40–42], information sheets from children’s hospitals [43, 44], and the Family Handbook™, a general guide to pediatric oncology [45]. The knowledge base was created to improve the accuracy of responses with evidence-based materials in-line with testing scenarios. Using natural language understanding (using large language model- GPT 3.5), the topic of interest was saved in a cache while content in the knowledge base was reformatted to summarize and simplify the original language based on the context. (3) Continuation: The user had an option to follow up on their initial inquiry, such as asking for more information about complications or asking for clarifications. At this state, the VA stored a copy of the conversation to regenerate a specific answer or expand with a comprehensive statement. Figure 1 outlines the VA prototype and interaction flow. Throughout the conversation, VA responses are extracted from a pediatric oncology guidebook as the knowledge base [45].

**Figure 1.**
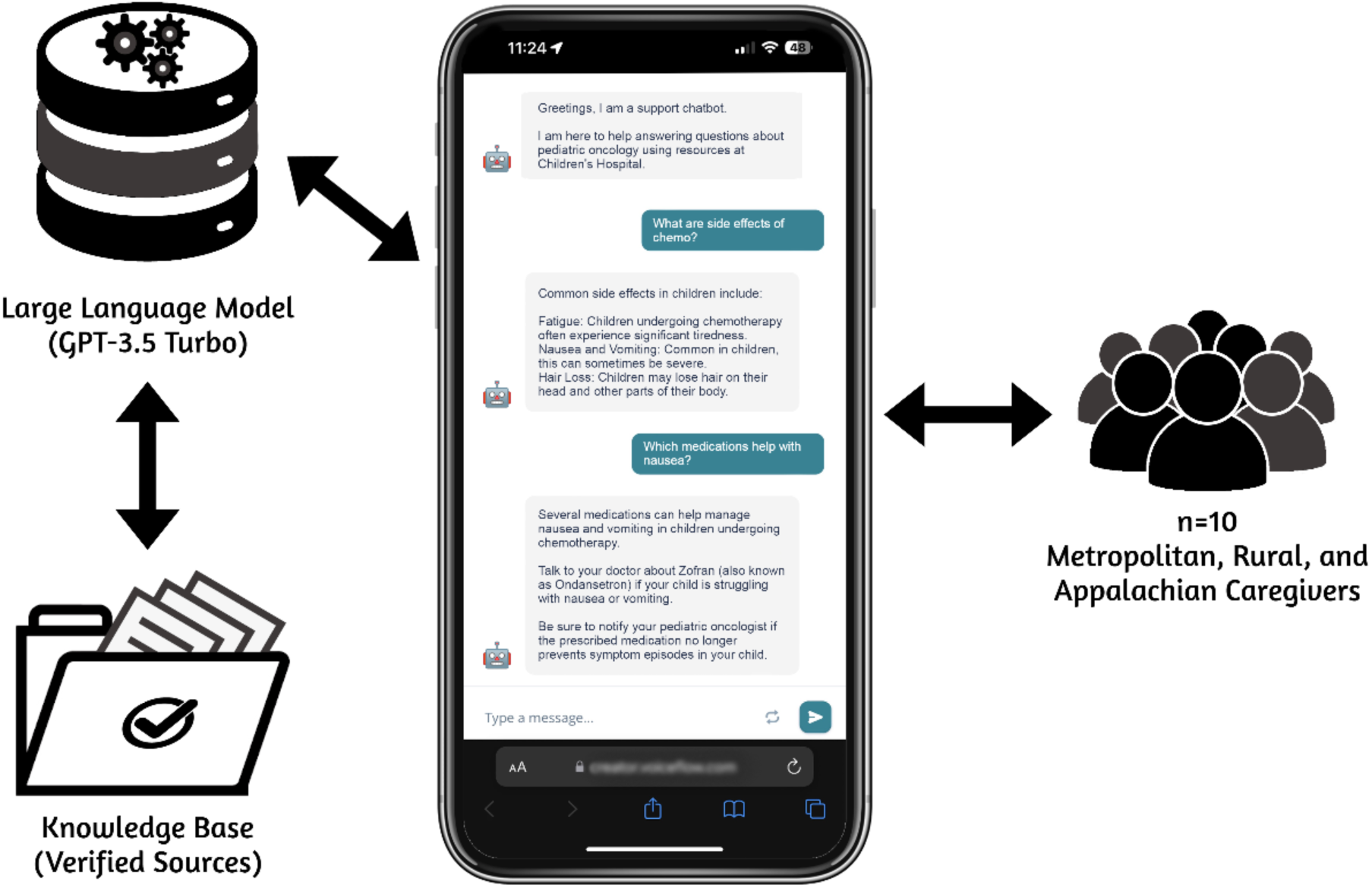
Flow diagram of the prototype web-based virtual assistant on a mobile smartphone.

### Sample Selection

Following IRB approval (#00002939), purposive sampling was employed at a large pediatric academic medical center to recruit caregivers of YCCS who had formerly participated in a stakeholder analysis (n=29) and/or were currently involved in a Community Advisory Board (*n* = 13; ∼40% rural or Appalachian) from a larger study. These groups involved caregivers of multiple types (biological mothers and fathers, grandparents, adoptive parents, and legal guardians) who had YCCS currently between the ages of 3 and 12 years, at least 6 months post-treatment or were on maintenance therapies, and without evidence of recurrent disease. We aimed to enroll 10 caregivers, which was in line with user experience studies in the literature [46, 47]. Eligible caregivers were defined as: (1) legal guardians, (2) living with their children ≥50% of the time, (3) residing in the surrounding states, and (4) English-speaking.

Caregivers who participated in interviews were compensated with a $50 gift card.

### Data Collection

We used a semi-structured qualitative interview methodology designed to capture insights from caregivers (See Appendix A for Interview Protocol). Ten participants were given the choice to participate in a focus group (n=7) or one-on-one interviews (n=3). The procedure began with an introduction to VA technology currently in use in daily life. Concepts related to popular market products (i.e., Siri from Apple, Google Assistant, & Alexa from Amazon) were briefly described based on their similar functionalities and potential. Study staff then asked caregivers to describe how they use VA-based technologies in their daily life.

Following the introduction, each caregiver interacted with a prototype VA for a set duration within the interview session. This phase included previewing 4 scenarios about late effects of cancer treatment, returning to school, reviewing cancer history, and handling the emotional impact on the child. This process guided caregivers’ engagement and helped them explore the VA’s features and functions thoroughly. Caregivers were also given opportunity to ask their own questions indirectly (queries monitored by study staff) to the prototype VA using voice or text. Finally, caregivers were prompted to express their thoughts about how a VA similar to the prototype could assist in their role as a caregiver to a YCCS.

After the engagement period, study staff solicited caregiver feedback on VA usability, utility, and feasibility. Focus groups and interviews lasted between 30 to 60 minutes (M=42, SD=13). All interactions were audio-recorded on Microsoft Teams with the caregivers’ consent and imported to NVivo.

Additionally, qualitative work incorporated Manning’s In Vivo [48] coding of transcription data through NVivo on release version 14.23.2. Recruitment for the study concluded after achieving data saturation within this niche pediatric oncology population. The research team determined that the current sample size provided rich and meaningful data for analysis. This approach aligns with the qualitative research in the literature [49].

### Data Analysis

Data was analyzed through a thematic analysis following Braun and Clarke’s [50] six-step process. Each dialogue file was cleaned, and study data was organized into an NVivo file. The conversations were divided into sentence segments, summarizing groups of sentences with code words derived from the dialogue. Co-authors [ES, DIJ] reviewed and coded the transcripts (Cohen’s Kappa, k=0.862). Conflicts were resolved through consensus discussion among authors. Subsequently, codes were organized into overarching themes.

## Results

### Demographics

The study sample consisted of caregivers of YCCS (n=10) who were predominantly female (n=9) with one male participant (n=1). In terms of relationship status, most were married (n=8), with one single caregiver (n=1) and another cohabitating (n=1). Educational attainment was high, with caregivers having at least a technical/trade school education (n=1), college degree (n=7), or graduate school education (n=1). One participant had a high school degree only (n=1). Annually, family income varied, with the highest earners making over $100,000 (n=3), followed by a few in the $75,001-$100,000 (n=2) and $50,001-$75,000 (n=3) ranges, and a couple earning between $25,000-$50,000 (n=1) or less than $25,001 (n=1). Most caregivers self-identified as White (n=9), with one participant identifying as Black (n=1).

Additionally, one participant endorsed having a Hispanic/Latino ethnicity (n=1). Child diagnoses were evenly distributed among leukemia/lymphoma (n=3), non-central nervous system solid tumors (n=4), and brain tumors (n=3). Child patients were distributed evenly since time of diagnosis (n=10 children, min=2.17 years, max=9.75 years, M=5.97 years, SD=2.80 years). Over half of children received one or more central nervous system-directed treatment (n=6), while the rest did not (n=4). Residential areas varied, with caregivers residing in metropolitan (n=4), rural (n=3), Appalachian (n=2), and rural- Appalachian (n=1) regions.

### Themes

Thematic analysis resulted in 22 codes and 4 themes: Interface and Interaction, User Experience, Content Relevance, and Trust (see Table 1). Interface and Interaction describes the different ways users preferred to access a VA, such as, through speech-only, text, or both. User Experience encompasses experiences and feelings about caregiving and seeking medical advice. Content Relevance describes the topics of value to caregivers and the accuracy of VA responses, such as, neurodevelopmental therapies, medication side effects, and educational resources. Trust describes security measures and the preference for confidentiality when handling VA data.

**Table 1:**
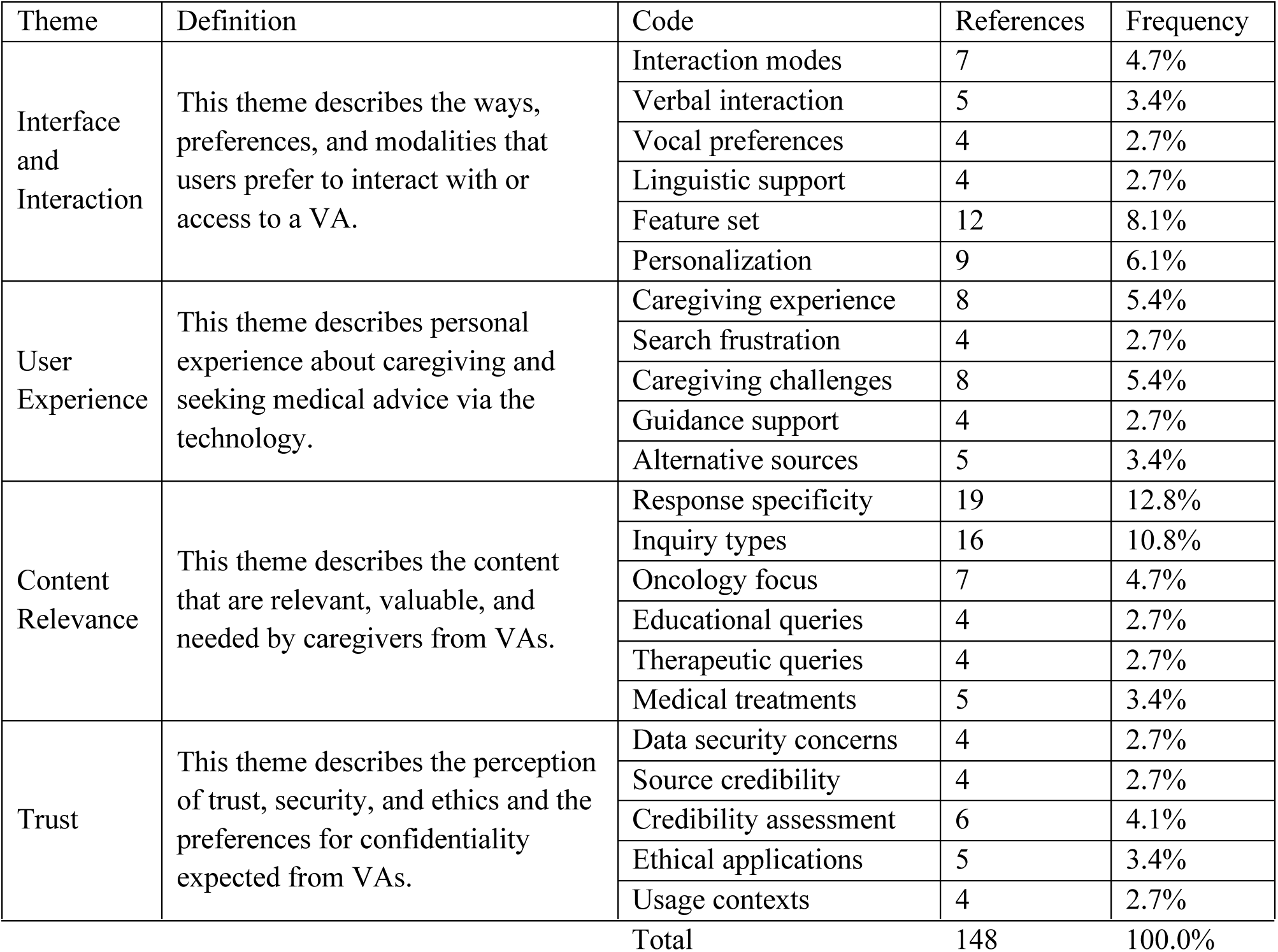
Themes and Codes.

#### 1. Interface and Interaction

Caregivers explained the technologies that they used in daily life, and their preferences for accessing and using VAs. They outlined that they have already used mHealth apps, search engines, and smart technologies to navigate the landscape of healthcare information, from connecting with their oncology team to managing a treatment plan long term. For some caregivers, VAs were a way to stay on top of daily life.

> *“Smart phones, you know the [smart speaker brand], the smart watches, all of that. I think helps [us] to keep a little organized.” Caregiver #10*

Caregivers also compared voice versus text-only interactions, and highlighted that the modality of conversations matter based on their location and situation. One caregiver described a scenario where voice interaction may not be appropriate for after-hours and text-based interaction could be more appropriate.

> *“When your child is ill and they’re finally asleep, you don’t wanna do anything to wake them up [so] you’re not gonna use voice.” Caregiver #7*

Caregivers acknowledged how VAs may handle administrative tasks well, such as reminders and scheduling, but struggle with extended conversations about medical information and questions. Caregivers desired to use VAs with a memory component (i.e., a knowledge base) for recalling past topics or exploring options at their healthcare center.

> *“What if I forgot what I had asked it, and I’m like, ‘Oh, there was some really good information there?’” Caregiver #1*
>
> *“There was something there that I wanted to revisit. So, maybe a search history within the [VA] app itself might be something valuable.” Caregiver #3*

Caregivers indicated that VAs could be integrated to the health system and referred by their clinicians for personalized oncological questions and responses. Many caregivers recognized how VAs for healthcare are typically on the landing page of a pediatric oncology clinic’s website to support navigating treatment services. Instead of using generic website VAs, several caregivers wanted to download an application to engage in further personal care questions, which was perceived to increase their access to information and convenience. Similarly, most caregivers noted the value of multi-language personal health education options. They indicated that most informational materials were printed and distributed in English-only.

One caregiver also mentioned having a VA feature that could educate users on common medical terminology.

> *“When you have a doctor’s visit, and they say, ‘oh, we’ve got this VA. Download the app, and you can ask your questions right there.” Caregiver #3*
>
> *“Could I ask it like a dictionary? Would you be able to ask things like, ‘what does hemoglobin mean?’ It would be great if I could.” Caregiver #5*“Unfortunately, there’s a very large portion of families that like, their ability to read is limited. So, if you can read back to them, you’re catching a whole other demographic that you’re missing when you provide pamphlets and stuff.” Caregiver #6

#### 2. User Experience

Most caregivers expressed that seeking health information has been challenging. Caregivers mentioned concern about “going down the rabbit hole” where they became stuck endlessly searching online. Many caregivers agreed that a major obstacle to accessing quality health information online was their difficulty finding useful medical advice specific to their child’s condition. Regardless of their backgrounds, caregivers were appreciative of the wealth of information that exists about oncology, yet acknowledged the limitations of navigating to that information and understanding it based on health literacy.

> *“We are given so much stuff and you just can’t keep track of everything.” Caregiver #1*
>
> *“[Do] you know what I mean? The medical terms that sometimes you’re given, and you have no clue, you know, what they’re talking about.” Caregiver #5*

Furthermore, caregivers recalled moments where they had to use alternative information sources (i.e., healthcare team); they would either take their concerns to their peer groups, or go through search engine results, acknowledging the risk of misinformation. Caregivers identified this cycle as a high contributor to caregiver burden and exhaustion. Alternatively, some caregivers were proactive with their network and contacted other healthcare providers for extra advice that is often not included in informational handouts and flyers.

> *“If Googling [an oncology symptom and diagnosis], there’s so much stuff out there… You don’t know what’s gonna come up!” Caregiver #2*
>
> *“As an oncology parent, if we have a quick question or we kind of need that ‘college of knowledge,’ we go to the [social media] page and get 50 answers right away.” Caregiver #10*
>
> *“I would try to find a couple nurses on the oncology floor saying, ‘Hey, you got a minute?’” Caregiver #1*
>
> *“It’s not just the doctors, but a lot of times the nurses have a lot more information to share about the day-to-day life.” Caregiver #4*

Caregivers summarized their needs from VA to access reliable answers quickly while filtering out less credible resources. Caregivers indicated VAs may reduce or eliminate both extended searching sessions and the burden paired with it in the future.

> *“This [the VA] would be a good resource for that tired, exhausted parent who’s at their wit’s end and just needs something.” Caregiver #7*

#### 3. Content Relevance

Caregivers expect VAs and other conversational technologies to deliver niche and highly accurate information. After interacting with the VA prototype, caregivers listed common topics of interest, such as educational and therapeutic information (e.g., learning more about speech therapy or special education options) to address neurodevelopmental delays and late effects, as well as treatment-based instructions.

While testing the prototype VA, caregivers reacted to the breadth of information it could cover, such as the use of therapeutics and mental health resources.

> *“Like, that’s a start. That is something where- you could take ‘that’ and go back to your physician and say… ‘Do we need occupational therapy to help them hold a pencil?’” Caregiver #1*
>
> *“Services for mental health of my pediatric cancer patient could be catered to provide information that [my] hospital would offer” Caregiver #10*
>
> *“You could ask it like, I’m driving from out of town, [are] there resources for me to stay [nearby]… while my child receives treatment.” Caregiver #6*

Caregivers expressed their use of VA with clarifying questions and scenario-based explanations. Some caregivers needed summaries, while others explored alternative interpretations of their doctor’s orders.

> *“…instead of having to cycle through the various pages to my messages or to my lab results, if I [could] just ask the question; ‘Hey, what was the summary of [patient’s] last [diagnostic procedure]?’” Caregiver #4*

Caregivers also referred to the need for evidence-based content and the VA’s ability to access credible resources, such as safe dosage instructions. They indicated that this information could be useful to determine when they should reach out to their oncology team:

> *“I would just say one of the first things that popped in my head when it comes to questions was definitely medication. I just think that would be, like very helpful to have that information here.” Caregiver #9*
>
> *“[The VA] made me think about how sometimes we get packets of information, and we don’t have the time to read through it. It would be great to have something like this to ask a question and get an answer right away.” Caregiver #8*
>
> *“It would be a great tool that people could use when it’s not necessarily worthy of calling a doctor, but they just need that additional resource.” Caregiver #7*

#### 4. Trust

Caregivers expressed that they expected their VAs to access credible sources and maintain confidentiality. Caregivers identified that it could be helpful to have a healthcare provider connected to the VA to monitor questions asked and provide urgent intervention when needed, rather than receiving strictly VA-based feedback. Caregivers also expressed that connecting a VA to a trusted provider or hospital system could also help them to trust the VA system and place more value on its answers. Finally, they expressed a potential point of frustration if the VA’s response or health advice comes from advertised or unreliable webpages:

> *“If you scroll at the very top… [reads opening message of prototype VA app], that helped me realize that they know this hospital and they’re aware of the resources that are available there…” Caregiver #6*
>
> *“…let them know like these are from trusted places; websites or facilities, studies… I think that will be very helpful because then it will put you guys separate from a general place we will go for other questions.” Caregiver #9*
>
> *“As long as they [VA answers] were [using] reliable sources, you know, like cancer organizations or making sure it’s like an org or something, or like the hospital websites versus… not like a [blog] answer.” Caregiver #7*

In line with that, endorsement or ownership of a VA by known health institutions was perceived to be a trust building indicator towards adopting, building trust, and using VA for personal health. Specifically, caregivers indicated the importance of displaying affiliations, logos, and titles from established organizations and healthcare systems.

> *“When you’re using this type of medium and you’re putting [hospital logo] on here, I’m going to immediately trust this as a reputable resource for my family’s medical inquiries.” Caregiver #4*
>
> *“If I have a dedicated resource that I know I trust by putting that name [points at hospital name & logo] on there, I think it [the VA] can go a long way.” Caregiver #8*

In terms of security, caregivers would rather log into an app or service than use publicly available VAs. Caregivers wanted the flexibility to discuss health-related problems without having to worry about how the conversational data is handled, highlighting the need for a trustworthy VA system.

> *“This [could] be like a secure kind of app that you could just like, log into once and then [use], versus [having] to log into [patient’s electronic health record portal].” Caregiver #10*

Caregivers also expressed that VAs need to use ethical practices, providing disclosures on its capabilities and knowledge expertise at the start of a conversation, and also indicating when a question warrants medical, rather than VA, consultation. Beyond low-risk question, such as benign symptoms, VAs were expected to clearly direct users when noticed as urgent cases. Additionally, some caregivers indicated that VAs could build trust via conversations, particularly in how it responds and guides high-risk cases:

> *“[My child] had bad cramps in [their] feet and legs. My first thought was, ‘Oh, is the cancer back?’ because it started from a limb. [sighs] I googled it and found it was actually very normal; they’re growing pains, so I just monitored it and know when to reach out to my physician.” Caregiver #1*
>
> *“’When should I consult my doctor about a fever?’ And then it could respond, ‘if the fever is 100.4 or more, you should contact your physician immediately’” Caregiver #10*

During the study, we observed slight differences in responses regarding residential groups (metropolitan, rural/ Appalachian, rural, Appalachian). Metropolitan caregivers focused mostly on access to information, documentation, and tracking treatment symptoms via technology. The rural and Appalachian caregivers focused mostly on the convenience and accessibility side of technology. We also observed that rural and Appalachian caregivers expressed more reliance on technology as a learning tool compared to Metropolitan caregivers to facilitate communication with their healthcare team.

## Discussion

Limited access to early intervention and related information for caregivers, particularly in rural and Appalachian areas, exacerbates recovery processes that could otherwise be mitigated with timely treatment. We investigated VA engagement with caregivers to explore the technology’s potential to bridge the knowledge accessibility gap while supporting marginalized communities. More specifically, this study provides critical insights into the perceptions of caregivers of young childhood cancer survivors regarding the use of virtual assistants for health information seeking and care management. We found that caregivers valued easily accessible platforms that could be adapted for multimodal engagement. The conversational design of VAs, such as tonal or visual features, was integral for caregivers validating their trust in VAs. The findings further emphasize the potential of VAs to address the information needs of these caregivers, particularly in underserved communities, by offering a convenient, personalized, and reliable source of information.

A common challenge expressed by caregivers was difficulty seeking and accessing reliable health information online, often experiencing frustration and exhaustion from extensive searching. Such an experience has other caregivers and patients using digital health technologies (e.g. mHealth apps, patient portals) [51]. Furthermore, difficulties navigating online health information are well-documented [52]. In our study, caregivers valued the potential of VAs to provide quick, and ideally reliable answers, thereby reducing their cognitive and emotional burden. Still, in the current state of commercial VAs, the literature reports mixed evidence on their utility and reliability [53]. Misinformation has been a risk factor, especially for online health information seeking [54, 55]. Therefore, the role of VAs in alleviating these challenges by filtering out less credible sources and providing immediate responses, provides an efficient means to access accurate and evidence-based information [56]. With a healthcare information database for pediatric oncology, which we developed as a proof-of-concept for this study, specialized VAs may have a role in the digital health ecosystem to improve the efficiency and effectiveness of health information seeking for caregivers [57, 58].

Different caregiver preferences regarding interaction with VAs (including speech-only, text-only, and a combination of both modalities) highlighted the expected flexibility from VAs to adapt to different contexts, such as using voice commands when hands-free interaction is needed or text-based interaction during quiet times (e.g., when the patient is resting). Adaptive modality to engage with digital health tools has been a common theme among caregivers of children with special healthcare needs [59, 60]. The ability of VAs to switch between modalities based on user context may further contribute to user experience and satisfaction [61, 62]. Therefore, the capabilities of VAs outlined in our study suggest the need for intelligent systems that support continuity in information seeking. To fit within the digital ecosystem of caregivers, such preferences should be integrated into VAs for care support. This finding is consistent with other populations with healthcare needs, who used conversational agents in patient care and remote care [63].

In line with caregiver preferences, the importance of personalization was highlighted, such as receiving specific, evidence-based content from VAs, particularly related to educational, therapeutic, neurodevelopmental, and treatment-based information. Earlier studies show alignment with our findings with other digital health technologies, such as mobile apps for cancer education, therapeutics and neurodevelopmental interventions [64, 65]. We found that these needs remain similar to those of other VAs. However, more personalization is required given the desire for highly specific and reliable health information, similar to studies on digital health interventions [66, 67].

Recent advancements in artificial intelligence and language models indicate an increasing ability for VAs to provide tailored information based on individual queries, which might be instrumental to address core needs for personalization [68].

Similar to user perceptions towards other digital health tools, trust and confidentiality has been an emerging concern, and therefore, it has been crucial to caregivers’ adoption of VAs [69, 70]. Caregivers expressed a preference for secure and confidential interactions, ideally through a dedicated system, which is consistent with literature on the use of patient portals [71, 72] and health information technology [73]. Moreover, the trust factor emerged as a multifaceted construct that identified expectations including data security, source credibility, and ethical implementations [74, 75]. Therefore, trust in VA may require further investigation.

Considering the residential differences, the literature suggests a potential disparity in digital health technology access and use among Rural and Appalachian populations [76–78]. Specifically, caregivers that live closer to major healthcare systems in metropolitan areas may have more opportunities to learn and organize general knowledge about oncology directly from their oncology specialists than those in rural and Appalachian regions who may have less access to specialized healthcare [79]. In our study with a limited sample size, we observed minimal differences among groups in their health-seeking attitudes and preferences for interacting with a VA. This suggests the need for further investigation of both groups to understand the needs and expectations of VA in patient care and how those might differ based on access to care.

### Clinical Implications

Our study suggests that caregivers of YCCS are largely open-minded and optimistic about the potential utility of VAs as they seek information and resources about their child’s oncology care, including treatment options and management of emerging late effects. Caregivers suggested in retrospect that, VAs may have been useful when their child was in active treatment to identify common side effects of medications or to track symptoms over time. Therefore, VAs might be an effective clinical communication tool for providers [29]. With future refinement and clinical validation, VAs might also facilitate early detection of late effects, such as prompting caregivers to track developmental milestones, which could then be flagged based on guidelines for further clinical assessment by a child’s healthcare provider. If medical or developmental needs are identified, VAs could also assist in streamlining referrals to relevant local resources based on where the family lives and waitlists, which may reduce provider and family burden. These advances could be particularly important in the context of families with limited access to integrated psychosocial and developmental care in less resourced areas, yet ongoing barriers to implementation, as previously identified, include liability, regulation, security, and privacy for VA and other technology solutions.

### Limitations

While this study provides insights into the perceptions of caregivers, several limitations must be acknowledged. One notable limitation is the potential bias in our sample. The caregivers who agreed to participate were part of an ongoing virtual Community Advisory Board (CAB) and may already be technologically inclined, which could skew the results towards a more favorable view of VAs. None of the caregivers expressed strong negative perceptions of VAs, which might not fully represent the broader population of caregivers who may be less familiar or uncomfortable with technology. Additionally, the sample included few caregivers from racially and ethnically marginalized backgrounds, limiting the generalizability of our findings across diverse populations. The study also focused solely on caregivers. Future work should gather healthcare provider perspectives regarding the utilization of VAs and directly test the clinical accuracy of the information provided with VAs in the pediatric oncology context. While qualitative methods are effective for exploring in-depth perceptions and experiences, they do not provide the same level of generalizability as quantitative or mixed method approaches. Furthermore, the potential for social desirability bias exists, as participants may have provided responses they believed were expected or favorable rather than their true feelings and experiences. The interaction period with the VA prototype was relatively short, which may have limited the depth of insights gained regarding long-term use and utility. Future work with extended interactions over a longer period could provide more detailed information on the practicality and effectiveness of VAs in supporting caregivers.

We presented preliminary evidence of the utility of VAs in supporting caregivers, and a preliminary understanding of the behaviors and needs of caregivers. These findings are relevant to policymakers and healthcare practitioners, enabling them to make informed decisions while incorporating VAs into health communication strategies, and understanding how perceptions might differ in these communities. This effort can lead to the effective tailoring of existing public health programs to integrate digital platforms. The preliminary findings from this study will further serve as a critical basis for future development and pilot testing of VA-based systems designed to meet the specialized needs of families of pediatric cancer patients and other clinical and chronic conditions.

## Conclusions

This study illuminates the potential of VAs to support caregivers of YCCS in navigating health information landscapes, particularly in underserved communities. By offering personalized, reliable, and easily accessible information, VAs can alleviate the cognitive and emotional burden on caregivers and improve their overall experience. Future research should focus on further refining VA functionalities and exploring their long-term impact on caregiver well-being and health outcomes.

## Data Availability

All de-identifiable data produced in the present study (interview codes and quotes) are available upon reasonable request to the authors.

## Statements and Declarations

### Competing Interests

All authors declare they have no financial nor non-financial competing interests.

### Ethics Approval

This study was performed in line with the principles of the National Institutes of Health. Approval was granted by the Institutional Review Board of Nationwide Children’s Hospital (#00002939).

### Consent to Participate

Informed consent was obtained from all individual participants included in the study.

## Notes

### Competing Interest Statement

The authors have declared no competing interest.

### Funding Statement

This study was funded by the intramural funding program of the Neurodevelopmental Research Affinity Group (NRAG) at Nationwide Children's Hospital. The Caregiver Advisory Board formation was supported by the American Psychological Association Division 54 Drotar-Crawford Postdoctoral Research Grant.

### Author Declarations

This study received ethical board approval by the Institutional Review Board of Nationwide Childrens Hospital (IRB# 00002939).

## References

1. Ostrom QT, Gittleman H, Xu J, et al. (2016) CBTRUS Statistical Report: Primary Brain and Other Central Nervous System Tumors Diagnosed in the United States in 2009-2013. Neuro Oncol 18:v1–v75. 10.1093/neuonc/now207

2. Phillips SM, Padgett LS, Leisenring WM, et al. (2015) Survivors of childhood cancer in the United States: prevalence and burden of morbidity. Cancer Epidemiol Biomarkers Prev 24:653–663. 10.1158/1055-9965.EPI-14-1418

3. Racine NM, Smith A, Pelletier W, et al (2018) Evaluation of a Support Group for Parents of Children Hospitalized for Cancer and Hematopoietic Stem Cell Transplantation. Soc Work Groups 41:276–290. 10.1080/01609513.2017.1356799

4. Sun C-L, Francisco L, Kawashima T, et al (2010) Prevalence and predictors of chronic health conditions after hematopoietic cell transplantation: a report from the Bone Marrow Transplant Survivor Study. Blood 116:3129–39; quiz 3377. 10.1182/blood-2009-06-229369

5. Wiener L, Kazak AE, Noll RB, et al (2015) Standards for the Psychosocial Care of Children With Cancer and Their Families: An Introduction to the Special Issue. Pediatr Blood Cancer 62 Suppl 5:S419–24. 10.1002/pbc.25675

6. National Academies of Sciences, Engineering, and Medicine, Institute of Medicine, Board on Health Care Services, National Cancer Policy Forum (2015) Comprehensive Cancer Care for Children and Their Families: Summary of a Joint Workshop by the Institute of Medicine and the American Cancer Society. National Academies Press

7. Marusak HA, Iadipaolo AS, Harper FW, et al. (2018) Neurodevelopmental consequences of pediatric cancer and its treatment: applying an early adversity framework to understanding cognitive, behavioral, and emotional outcomes. Neuropsychol Rev 28:123–175. 10.1007/s11065-017-9365-1

8. Willard VW, Cox LE, Russell KM, et al (2017) Cognitive and psychosocial functioning of preschool-aged children with cancer. J Dev Behav Pediatr 38:638–645. 10.1097/DBP.0000000000000512

9. Grieco J, Evans CL, Yock T, Pulsifer M (2022) Early psychosocial and executive functioning outcomes in pediatric brain tumor survivors after proton radiation

10. Jacola LM, Partanen M, Lemiere J, et al (2021) Assessment and monitoring of neurocognitive function in pediatric cancer. J Clin Oncol 39:1696–1704. 10.1200/jco.20.02444

11. Cox LE, Kenney AE, Harman JL, et al (2019) Psychosocial functioning of young children treated for cancer: Findings from a clinical sample. J Pediatr Oncol Nurs 36:17–23. 10.1177/1043454218813905

12. National Academies of Sciences, Engineering, and Medicine, Health and Medicine Division, Board on Health Care Services, National Cancer Policy Forum (2018) Long-Term Survivorship Care After Cancer Treatment: Proceedings of a Workshop. National Academies Press

13. Harman JL, Wise J, Willard VW (2018) Early intervention for infants and toddlers: Applications for pediatric oncology. Pediatr Blood Cancer 65:. 10.1002/pbc.26921

14. Yao N, Alcalá HE, Anderson R, Balkrishnan R (2017) Cancer Disparities in Rural Appalachia: Incidence, Early Detection, and Survivorship. J Rural Health 33:375–381. 10.1111/jrh.12213

15. Fish M, Pinkerman B (2003) Language skills in low-SES rural Appalachian children: normative development and individual differences, infancy to preschool. J Appl Dev Psychol 23:539–565. 10.1016/S0193-3973(02)00141-7

16. Rhoad-Drogalis A, Justice LM (2018) Absenteeism in Appalachian preschool classrooms and children’s academic achievement. J Appl Dev Psychol 58:1–8. 10.1016/j.appdev.2018.07.004

17. Shorey S, Lau LST, Tan JX, et al (2021) Families With Children With Neurodevelopmental Disorders During COVID-19: A Scoping Review. J Pediatr Psychol 46:514–525. 10.1093/jpepsy/jsab029

18. Lambert SD, Loiselle CG (2007) Health information seeking behavior. Qual Health Res 17:1006– 1019. 10.1177/1049732307305199

19. Bright MA, Fleisher L, Thomsen C, et al (2005) Exploring e-Health usage and interest among cancer information service users: the need for personalized interactions and multiple channels remains. J Health Commun 10 Suppl 1:35–52. 10.1080/10810730500265609

20. Luger E, Moran S, Rodden T (2013) Consent for all: revealing the hidden complexity of terms and conditions. Association for Computing Machinery

21. Myers C, Furqan A, Nebolsky J, et al (2018) Patterns for How Users Overcome Obstacles in Voice User Interfaces. Association for Computing Machinery

22. Yang S, Lee J, Sezgin E, et al (2021) Clinical Advice by Voice Assistants on Postpartum Depression: Cross-Sectional Investigation Using Apple Siri, Amazon Alexa, Google Assistant, and Microsoft Cortana. JMIR Mhealth Uhealth 9:e24045. 10.2196/24045

23. Sezgin E, Militello LK, Huang Y, Lin S (2020) A scoping review of patient-facing, behavioral health interventions with voice assistant technology targeting self-management and healthy lifestyle behaviors. Transl Behav Med 10:606–628. 10.1093/tbm/ibz141

24. Sezgin E, Huang Y, Ramtekkar U, Lin S (2020) Readiness for voice assistants to support healthcare delivery during a health crisis and pandemic. NPJ Digit Med 3:122. 10.1038/s41746-020-00332-0

25. Amershi S, Cakmak M, Knox WB, Kulesza T (2014) Power to the People: The Role of Humans in Interactive Machine Learning. AIMag 35:105–120. 10.1609/aimag.v35i4.2513

26. Brewer RN, Findlater L, Kaye J “jofish,”, et al (2018) Accessible Voice Interfaces. Association for Computing Machinery

27. Nobles AL, Leas EC, Caputi TL, et al (2020) Responses to addiction help-seeking from Alexa, Siri, Google Assistant, Cortana, and Bixby intelligent virtual assistants. NPJ Digit Med 3:11. 10.1038/s41746-019-0215-9

28. Wagner D, Churchill A, Sigtia S, et al (2024) A multimodal approach to device-directed speech detection with large language models. In: ICASSP 2024 - 2024 IEEE International Conference on Acoustics, Speech and Signal Processing (ICASSP). IEEE, pp 10451–10455

29. Sezgin E (2024) Redefining virtual assistants in health care: The future with large language models. J Med Internet Res 26:e53225. 10.2196/53225

30. Vaidyam AN, Wisniewski H, Halamka JD, et al (2019) Chatbots and Conversational Agents in Mental Health: A Review of the Psychiatric Landscape. Can J Psychiatry 64:456–464. 10.1177/0706743719828977

31. Xu L, Sanders L, Li K, Chow JCL (2021) Chatbot for Health Care and Oncology Applications Using Artificial Intelligence and Machine Learning: Systematic Review. JMIR Cancer 7:e27850. 10.2196/27850

32. Chaix B, Bibault J-E, Pienkowski A, et al (2019) When Chatbots Meet Patients: One-Year Prospective Study of Conversations Between Patients With Breast Cancer and a Chatbot. JMIR Cancer 5:e12856. 10.2196/12856

33. Sezgin E, Hussain S-A, Rust S, Huang Y (2023) Extracting Medical Information From Free-Text and Unstructured Patient-Generated Health Data Using Natural Language Processing Methods: Feasibility Study With Real-world Data. JMIR Form Res 7:e43014. 10.2196/43014

34. Anderson JQ, Rainie H (2014) The Internet of Things Will Thrive by 2025. Pew Research Center [Internet & American Life Project]

35. Catania F, Spitale M, Garzotto F (2023) Conversational Agents in Therapeutic Interventions for Neurodevelopmental Disorders: A Survey. ACM Comput Surv 55:1–34. 10.1145/3564269

36. Creswell JW, Creswell JD (2022) Research design, 6th ed. Sage Publications, Christchurch, New Zealand

37. Voiceflow. https://www.voiceflow.com/. Accessed 24 Jul 2024

38. OCECD Publications. https://www.ocecd.org/Publications1.aspx. Accessed 24 Jul 2024

39. (2018) PEER SUPPORT MATTERS. In: Momcology. https://momcology.org-stg.gmheventweb.com/peer-support-matters/. Accessed 24 Jul 2024

40. Sitemap. https://education.ohio.gov/Miscellaneous/Sitemap. Accessed 24 Jul 2024

41. Resources and Publications. https://ddc.ohio.gov/resources-and-publications. Accessed 24 Jul 2024

42. (2017) Individuals with Disabilities Education Act (IDEA). In: Individuals with Disabilities Education Act. https://sites.ed.gov/idea/. Accessed 24 Jul 2024

43. Medicines Used to Treat Pediatric Cancer. In: St. Jude together. https://together.stjude.org/en-us/diagnosis-treatment/medicines-list.html. Accessed 24 Jul 2024

44. Cancer Resources from OncoLink, Treatment, Research, Coping, Clinical Trials, Prevention OncoLink Tools in PennChart. https://www.oncolink.org/oncolink-tools-in-pennchart. Accessed 24 Jul 2024

45. Children’s Oncology Group (2023) Children’s Oncology Group Family Handbook for children with cancer, 2nd ed. St. Baldrick’s Foundation

46. Motta M, Groves E, Schneider A, et al (2023) Designing self-tracking experiences: A qualitative study of the perceptions of barriers and facilitators to adopting digital health technology for automatic urine analysis at home. PLOS Digit Health 2:e0000319. 10.1371/journal.pdig.0000319

47. Tilley DO, McKeon B, Ibrahim N, et al (2024) A snapshot on a journey from frustration to readiness-A qualitative pre-implementation exploration of readiness for technology adoption in Public Health Protection in Ireland. PLOS Digit Health 3:e0000453. 10.1371/journal.pdig.0000453

48. Manning J (2017) In Vivo Coding. The International Encyclopedia of Communication Research Methods 1–2

49. Braun V, Clarke V (2021) To saturate or not to saturate? Questioning data saturation as a useful concept for thematic analysis and sample-size rationales. Qual Res Sport Exerc Health 13:201–216. 10.1080/2159676x.2019.1704846

50. Braun V, Clarke V (2006) Using thematic analysis in psychology. Qual Res Psychol 3:77–101. 10.1191/1478088706qp063oa

51. Choukou M-A, Olatoye F, Urbanowski R, et al (2023) Digital health technology to support health care professionals and family caregivers caring for patients with cognitive impairment: Scoping review. JMIR Ment Health 10:e40330. 10.2196/40330

52. Strand T, Westergren T (2024) A meta-ethnography on parents’ experiences of the Internet as a source of health information. Glob Qual Nurs Res 11:. 10.1177/23333936241259246

53. Alagha EC, Helbing RR (2019) Evaluating the quality of voice assistants’ responses to consumer health questions about vaccines: an exploratory comparison of Alexa, Google Assistant and Siri. BMJ Health Care Inform 26:e100075. 10.1136/bmjhci-2019-100075

54. Fridman I, Johnson S, Elston Lafata J (2023) Health information and misinformation: A framework to guide research and practice. JMIR Med Educ 9:e38687. 10.2196/38687

55. Wang Y, McKee M, Torbica A, Stuckler D (2019) Systematic literature review on the spread of health-related misinformation on social media. Soc Sci Med 240:112552. 10.1016/j.socscimed.2019.112552

56. Dutsinma FLI, Pal D, Funilkul S, Chan JH (2022) A systematic review of voice assistant usability: An ISO 9241–11 approach. SN Comput Sci 3:. 10.1007/s42979-022-01172-3

57. Kumah-Crystal Y, Pirtle C, Whyte H, et al (2018) Electronic health record interactions through voice: A review. Appl Clin Inform 09:541–552. 10.1055/s-0038-1666844

58. Hong G, Folcarelli A, Less J, et al (2021) Voice assistants and cancer screening: A comparison of Alexa, Siri, Google assistant, and Cortana. Ann Fam Med 19:447–449. 10.1370/afm.2713

59. Sezgin E, Noritz G, Lin S, Huang Y (2021) Feasibility of a voice-enabled medical diary app (SpeakHealth) for caregivers of children with special health care needs and health care providers: Mixed methods study. JMIR Form Res 5:e25503. 10.2196/25503

60. Sezgin E, Weiler M, Weiler A, Lin S (2018) Proposing an ecosystem of digital health solutions for teens with chronic conditions transitioning to self-management and independence: Exploratory qualitative study. J Med Internet Res 20:e10285. 10.2196/10285

61. Kadam AV (2022) Crafting Multi-Modal Interactions on Voice Assistants. Journal of Engineering and Applied Sciences Technology 4:1–5

62. Lee J, Wang J, Brown E, et al (2024) GazePointAR: A context-aware multimodal voice assistant for pronoun disambiguation in wearable augmented reality. In: Proceedings of the CHI Conference on Human Factors in Computing Systems. ACM, New York, NY, USA

63. Tennant R, Allana S, Mercer K, Burns CM (2022) Caregiver expectations of interfacing with voice assistants to support complex home care: Mixed methods study. JMIR Hum Factors 9:e37688. 10.2196/37688

64. Denis F, Maurier L, Carillo K, et al (2022) Early detection of neurodevelopmental disorders of toddlers and postnatal depression by mobile health app: Observational cross-sectional study. JMIR MHealth UHealth 10:e38181. 10.2196/38181

65. Khan K, Hall CL, Davies EB, et al (2019) The effectiveness of Web-based interventions delivered to children and young people with neurodevelopmental disorders: Systematic review and meta-analysis. J Med Internet Res 21:e13478. 10.2196/13478

66. Erku D, Khatri R, Endalamaw A, et al (2023) Digital health interventions to improve access to and quality of primary health care services: A scoping review. Int J Environ Res Public Health 20:. 10.3390/ijerph20196854

67. Wienert J, Jahnel T, Maaß L (2022) What are digital public health interventions? First steps toward a definition and an intervention classification framework. J Med Internet Res 24:e31921. 10.2196/31921

68. Chen J, Liu Z, Huang X, et al (2024) When large language models meet personalization: perspectives of challenges and opportunities. World Wide Web 27:1–45. 10.1007/s11280-024-01276-1

69. Bolton T, Dargahi T, Belguith S, et al (2021) On the security and privacy challenges of virtual assistants. Sensors (Basel) 21:2312. 10.3390/s21072312

70. Security and Privacy Concerns in The Implementation of Virtual Assistants in Multinational Companies

71. Latulipe C, Mazumder SF, Wilson RKW, et al (2020) Security and privacy risks associated with adult patient portal accounts in US hospitals. JAMA Intern Med 180:845–849. 10.1001/jamainternmed.2020.0515

72. Sethness JL, Golub S, Evans YN (2023) Adolescent patient portals and concerns about confidentiality. Curr Opin Pediatr 35:430–435. 10.1097/MOP.0000000000001252

73. Egan KJ, Clark P, Deen Z, et al (2022) Understanding current needs and future expectations of informal caregivers for technology to support health and well-being: National survey study. JMIR Aging 5:e15413. 10.2196/15413

74. Manzini A, Keeling G, Marchal N, et al (2024) Should users trust advanced AI assistants? Justified trust as a function of competence and alignment. In: The 2024 ACM Conference on Fairness, Accountability, and Transparency. ACM, New York, NY, USA

75. Wienrich C, Reitelbach C, Carolus A (2021) The trustworthiness of voice assistants in the context of healthcare investigating the effect of perceived expertise on the trustworthiness of voice assistants, providers, data receivers, and automatic speech recognition. Front Comput Sci 3:. 10.3389/fcomp.2021.685250

76. Okobi E, Adigun AO, Ozobokeme O-E, et al (2023) Examining disparities in ownership and use of digital health technology between rural and urban adults in the US: An analysis of the 2019 Health Information National Trends Survey. Cureus 15:e38417. 10.7759/cureus.38417

77. Tudor HL, Ingram R, Wackerbarth S (2023) Patient engagement in patient portals in Appalachia v. Surrounding U.s. census regions: An analysis of HINTS (health information national trends survey) data, 2017-2020. J Appalach Health 5:50–65. 10.13023/jah.0502.05

78. Vanderpool RC, Stradtman LR, Gaysynsky A, et al (2021) Access to and use of technology for health. 3

79. Kent EE, Lee S, Asad S, et al (2023) “If I wasn’t in a rural area, I would definitely have more support”: social needs identified by rural cancer caregivers and hospital staff. J Psychosoc Oncol 41:393–410. 10.1080/07347332.2022.2129547

